# Dominant extrafollicular B cell responses in severe COVID-19 disease correlate with robust viral-specific antibody production but poor clinical outcomes

**DOI:** 10.1101/2020.04.29.20083717

**Authors:** Matthew C. Woodruff, Richard P. Ramonell, Kevin S. Cashman, Doan C. Nguyen, Ankur Singh Saini, Natalie Haddad, Ariel M. Ley, Shuya Kyu, J. Christina Howell, Tugba Ozturk, Saeyun Lee, Weirong Chen, Jacob Estrada, Andrea Morrison-Porter, Andrew Derrico, Fabliha A. Anam, Monika Sharma, Henry Wu, Sang N. Le, Scott A. Jenks, Christopher M. Tipton, John L. Daiss, William T. Hu, F. Eun-Hyung Lee, Ignacio Sanz

## Abstract

A wide clinical spectrum has become a hallmark of the *SARS-CoV-2* (COVID-19) pandemic, although its immunologic underpinnings remain to be defined. We have performed deep characterization of B cell responses through high-dimensional flow cytometry to reveal substantial heterogeneity in both effector and immature populations. More notably, critically ill patients displayed hallmarks of extrafollicular B cell activation as previously described in autoimmune settings. Extrafollicular activation correlated strongly with large antibody secreting cell expansion and early production of high levels of *SARS-CoV-2*-specific antibodies. Yet, these patients fared poorly with elevated inflammatory biomarkers, multi-organ failure, and death. Combined, the findings strongly indicate a major pathogenic role for immune activation in subsets of COVID-19 patients. Our study suggests that, as in autoimmunity, targeted immunomodulatory therapy may be beneficial in specific patient subpopulations that can be identified by careful immune profiling.

## Main Text

As efforts intensify to identify treatment options and develop effective vaccination strategies against COVID-19 infection ^1, 2, 3, 4^, there is an urgent need for accurate characterization of protective and non-protective B-cell responses. The identification of new human B cell subsets ^5, 6, 7, 8^, and their integration in a coherent classification ^8^, enable the application of high-dimensional flow cytometry to accurately identify B cell profiles while providing functional context in order to understand infection response course. To this end, we studied 17 patients with confirmed COVID-19 infection with flow panels that: 1. Accurately identify B cell populations; 2. Assess activation status; and 3. Indicate homing potential through integrin and chemokine receptor modulation (Supplemental table 1). Of the COVID-19 cohort, 10 were critically ill requiring ICU admission (Severe COVID-19; ICU-C) and 4 of the 10 patients died (Supplementary table 2). This group was compared to milder outpatients (Outpatient COVID; OUT-C) and healthy donors (HD).

Despite reports of lymphocytopenia ^9^, patients with COVID-19 displayed elevated numbers of peripheral blood mononuclear cells, with CD19+ B cells significantly increased relative to HD (Supplemental fig 1). B cell profiling identified primary populations (transitional [**Tr**], naive [**N**], double negative [**DN**], memory [**M**], and antibody secreting cells [**ASCs**]), that could be then fractionated into 14 non-redundant, secondary B cell populations of established significance (supplemental Table 3) ^10^. As illustrated in Fig 1a-f, the three clinical groups display characteristic B cell profiles characterized by either expansions of ASC and DN2 cells (ICU-C) or transitional cells (OUT-C) (Figs 1-3).

**Figure 1.**
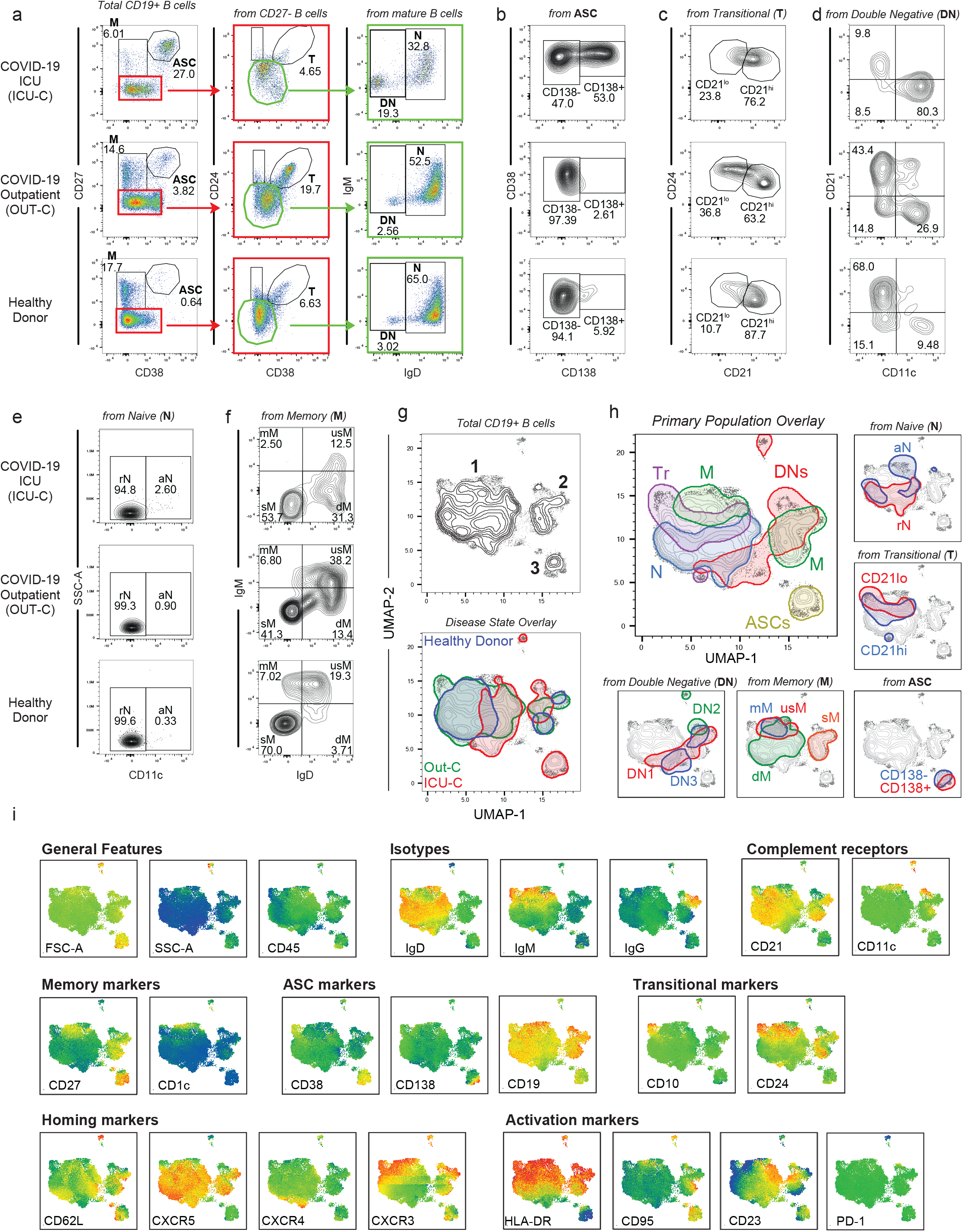
B cell characterization in acute COVID-19 infection. (a-f) Peripheral blood mononuclear cells (PBMCs) were isolated from healthy donors, or PCR+ SARS-CoV-2 outpatient, or ICU admitted patient bloods. Isolated cells were stained and analyzed by flow cytometry. (a) Primary population gating of representative patient samples. M - Memory B cells; ASC - Antibody secreting cells; Tr - Transitional B cells; DN - Double negative B cells; N - Naive B cells (b) Antibody secreting cell sub-gating (CD138+ ASC, CD138-ASC) of representative patient samples displaying an increase in CD138+ ASCs in ICU-C. (c) Transitional B cell sub-gating (CD21^lo^ Tr, CD21^hi^ Tr) of representative patient samples displaying an increase in CD21^lo^ Tr in OUT-C. (d) Double negative B cell sub-gating (Top left - DN1; Bottom left - DN3; Bottom right - DN2) of representative patient samples displaying an increase in DN2s in ICU-C. (e) Naïve B cell sub-gating (rN, aN) of representative patient samples. (f) Memory B cell sub-gating (mM, usM, dM, sM) of representative patient samples displaying a decrease in usM in ICU-C. (g) Top – UMAP projection of composite patient sample. Composite derived from 1000 representative cells from all analyzed patients. Bottom – Patient disease status overlaid on composite UMAP projection. (h) Primary and secondary populations as gated in [1a-f] overlaid on composite UMAP projection. (i) Heatmaps of select marker expression overlaid on composite UMAP projection. (g-i) Outlined regions contain 90% of cells derived from the indicated classification.

To further guide exploration of the cohort, a UMAP algorithm for dimensionality reduction ^11^ was applied to a cohort composite sample for all assessed parameters, and cells from the composite were mapped in 2-dimensional Cartesian space (Fig 1h). This approach reveals clear phenotypic B cell separation in the 3 clinical groups (Fig 1h). Thus, three major cell clusters were apparent in the projection. Cluster 1 is highly diverse, predominantly composed of transitional, naive, and IgM+ memory subsets. (Fig 1h). If note, while IgD-only memory (dM) is relatively rare in HD (Fig 1f), its diverse in surface phenotype resulted in a wide distribution across Cluster 1 (Fig 1h). Cluster 2 is more homogenous, consisting of DN populations 1-3, and the switch memory (sM) compartment. Importantly, sM B cells neatly overlay the DN1 population confirming our previous report of their close phenotypic association ^6^ while DN2 B cells formed a distinct cluster within the larger DN density (Fig 1i). ASC populations split by CD138 expression pattern defined cluster 3 (Fig 1i)

By overlaying disease states onto the composite and subtracting overlapping densities, differential cellular signatures become visually apparent (Fig 2a), with three main regions of interest distinguishing ICU-C and OUT-C patients both from each other, and from HD. Region 1 highlights an area of high CD11c expression by the ICU-C cohort demarcating activated-naive (aN) and DN2 populations (Fig 1i, Fig 2b), which are significantly enriched in ICU-C (Fig 2c-d). In accordance with previous studies defining their participation in EF responses ^8^ (Supplemental figure 2), both aN and DN2 cells express the highest levels of CD11c and of the IFN*γ*-inducible transcription factor T-bet (Fig 2e-f). Consistent with a critical role of similar B cells in autoimmunity and viral clearance in mice ^12, 13, 14^, our findings provide original evidence for the triggering of these B cells in human acute viral infection and in particular, in the early stages of COVID-19 infection. Region 2 is comprised of ASCs, a population whose expansion is a major correlate of protection in vaccine responses ^15^. In contrast, in COVID-19 infection, high levels of ASC formation instead portended negative disease outcomes with ICU-C patients presenting with significantly higher levels of ASCs than other groups (Fig 2g-h). These findings are consistent with a proposed immunopathogenic role of plasmablasts or circulating ASC expansions in acute Dengue infection ^16^. Similar to some vaccine responses ^17^, an unusually high ASC fraction expressed the plasma cell maturation marker CD138 (Fig 2i-j). This feature is likely to reflect the intensity and inflammatory milieu of the ASC response in ICU-C patients. Finally, region 3 maps neatly to the CD21^lo^ Tr subset (Fig 1h, Fig 2a), which is significantly increased in OUT-C patients. Transitional cells in general comprise up to 25% of the overall B cell compartment in OUT-C patients with CD21^lo^ cells making up more than half of the overall frequency gain (Fig 2k-l). Notably, while OUT-C CD21^lo^ Tr cells express similar levels of developmental markers such as CD10 and CD24 as HD (Fig 2m), they diverge in the expression of CD138 expression. Their correlation with milder COVID-19 disease is consistent with a protective role possibly played through regulatory functions ^18^. Alternatively, their higher expression of CD138 might be consistent with effector conversion into ASC as it has been recently described ^19^ (Fig 2m). Whether CD21lo Tr cells play a protective role through regulatory functions or the production of protective anti-viral antibodies should be elucidated in future studies.

**Figure 2.**
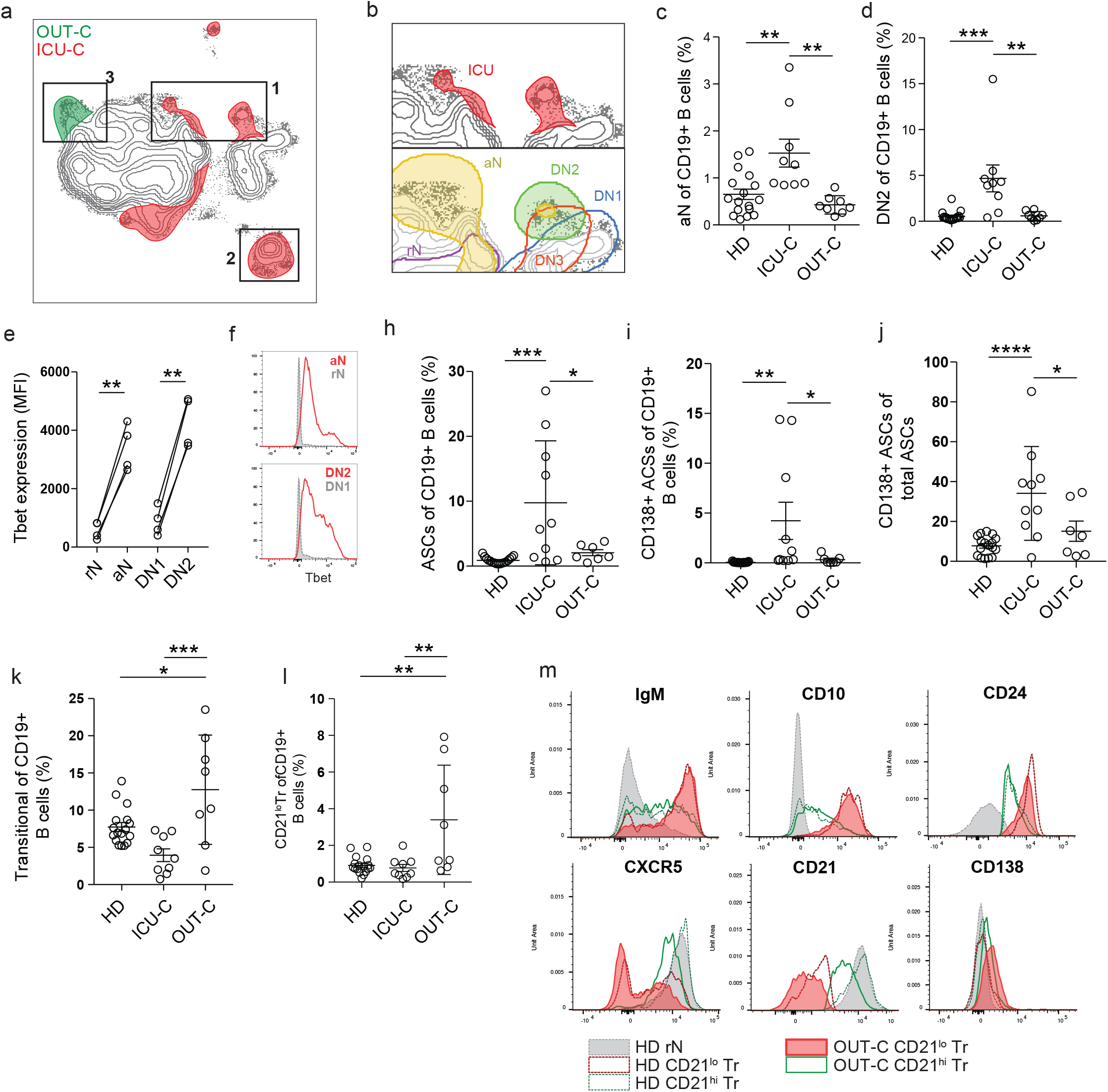
Unique B cell utilization by COVID-19 ICU and outpatients. (a) Overlay of patient disease status on composite UMAP projection as in [1g]. Regions of overlapping density are subtracted to display regions of unique population use. (b) Top – Magnification of region 1 from [2a]. Bottom – Indicated secondary populations overlaid on magnification of region 1 from [2a]. (c) aN frequency of CD19+ B cells in HD, UCI, or outpatients. (d) DN2 frequency of CD19+ B cells in HD, UCI, or outpatients. (e) T-bet expression in indicated secondary populations from COVID-19 ICU patients through intracellular staining and flow cytometry. (f) Representative histograms of T-bet expression as in [2e]. (g) ASC frequency of CD19+ B cells in HD, UCI, or outpatients. (h) CD138+ ASC frequency of CD19+ B cells in HD, UCI, or outpatients. (i) CD138+ ASC frequency of total ASCs in HD, UCI, or outpatients. (i) Tr frequency of CD19+ B cells in HD, UCI, or outpatients. (j) CD21^lo^ Tr frequency of CD19+ B cells in HD, ICU, or outpatients. (k) Histograms of indicated marker expression by flow cytometry.

The use of secondary population frequencies as features for the hierarchical clustering of COVID-19 patients, demonstrates a distinct separation between outpatient and ICU-admitted patients with COVID-19, driven primarily by a coordinated increase in the more severe group of both aN and DN2 populations, both EF pathway constituents (Fig 3a). A novel DN population (DN3), previously unreported in other conditions and defined by the absence of both CD21 and CD11c, consistently fell alongside aN and DN2 populations. While the function and significance of DN3 cells remain to be fully elucidated, these cells contain a CD38+ subset that is expanded in some patients, and correspond to the early pre-ASCs we previously identified in vaccine responses ^20^ (Supplemental Figure 2a). Their expansion in the ICU-C cohort, association with other EF populations, and reduced expression of CXCR5 suggests relevance to this effector pathway (Fig 3a, Supplemental Figure 2b-c). Expression of all three of these populations was highly correlated with the ASC expansion described above (Fig 2a, 3a), forming a clear EF population cluster that defined the responses within the critically ill. This was not a reflection of infection duration as both EF-high (CoV-A) and EF-low (CoV-B) clusters were sampled at similar times post symptom onset (Fig 3b).

**Figure 3.**
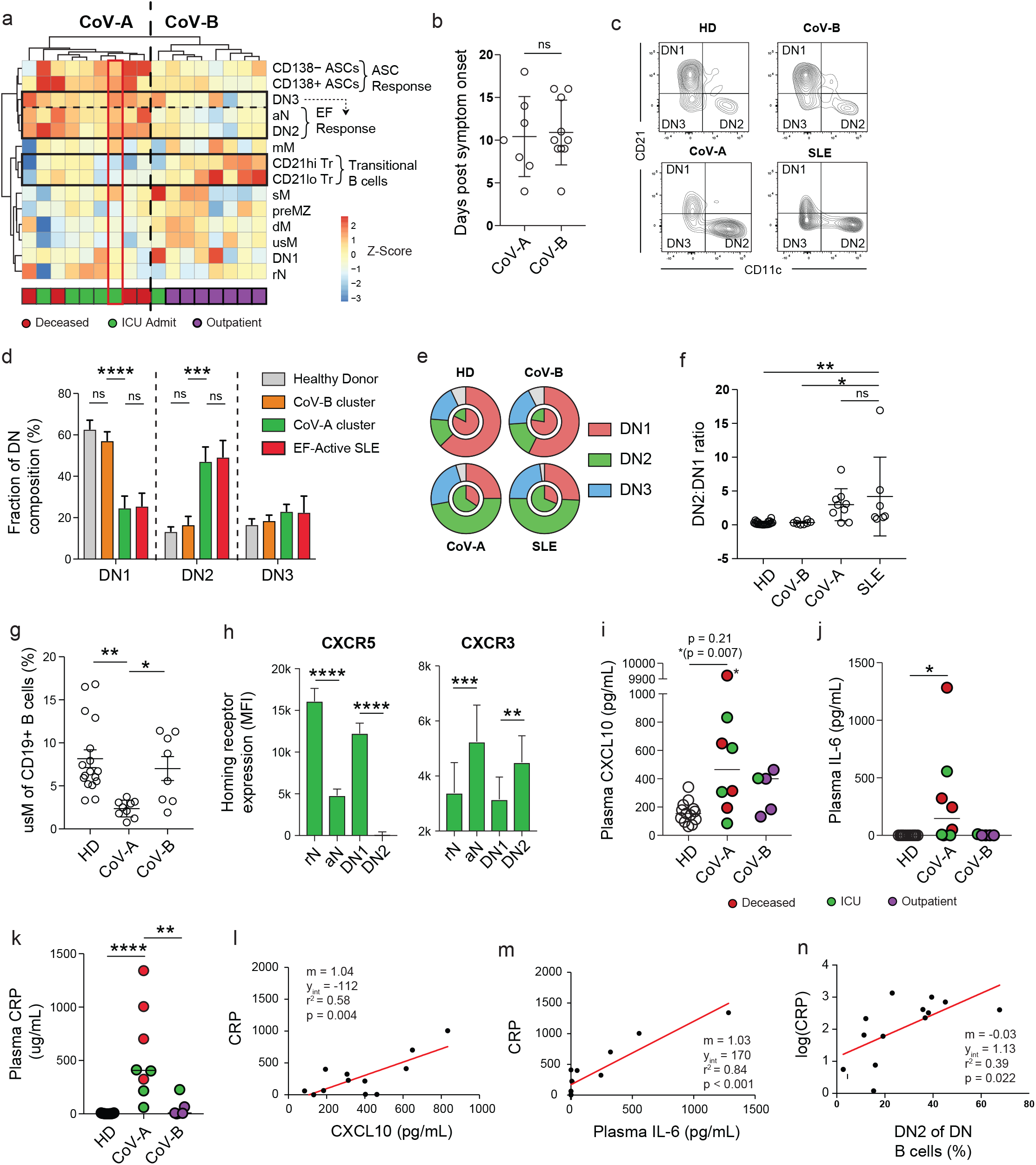
Extrafollicular response intensity is correlated with COVID-19 disease severity. (a) Heatmap of secondary population frequency z-scores by outpatient (blue), ICU admitted (green), or deceased (red) COVID-19+ patients. Associated dendrograms represent hierarchical clustering of patient samples and B cell populations by Ward’s method. Clusters designated as CoV-A and CoV-B for downstream analysis. Red box indicates patient analyzed by single cell VDJ analysis in Figure 4. (b) Patient sample collection times following symptom onset in CoV-A and CoV-B clusters. (c) Representative plots of DN population composition in HD, CoV-A, CoV-B, and SLE patient groups. (d) DN composition analysis in HD, CoV-A, CoV-B, and SLE patient groups. (e) Outer ring – Mean DN population composition of patient groups. Inner ring – Mean DN2 : DN1 ratios of patient groups. (f) DN2 : DN1 ratios in HD, CoV-A, CoV-B, and SLE patient groups. (g) usM frequency of CD19+ B cells in HD, CoV-A, or CoV-B groups. (h) Homing receptor surface expression in follicular (rN, DN1) vs. extrafollicular (aN, DN2) populations observed in CoV-A patients. (i) IP-10 concentration of HD, CoV-A, or CoV-B patient group plasma. Highest value (*) exceeded the testing range and was identified to be a statistical outlier (p < 0.00001). p-value reporting in parentheses indicates testing with outlier removed (j) IL-6 concentration of HD, CoV-A, or CoV-B patient group plasma (k) C-reactive protein (CRP) concentration of HD, CoV-A, or CoV-B patient group plasma (l) Linear regression of CRP values as a function of IP-10 plasma levels (with outlier removed). (m) Linear regression of CRP values as a function of IL-6 plasma levels. (n) Linear regression of log(CRP) values as a function of DN2 B cell frequency of total DN B cells. (b) Student’s t testing displayed between groups. (c-j) ANOVA analysis with Tukey’s multiple comparisons testing between groups. (c-n) *p <= 0.05; **p <= 0.01; ***p <= 0.001; **** p <= 0.0001.

The combination of T-bet driven effector development and high frequency of ASC in ICU-C patients is highly similar to previous studies of the activation of the EF B cell pathway in patients with highly active SLE^6^. In those studies, activation of the B cell EF pathway resulted in expansions in the ASC, aN, and DN2 compartments with concordant reductions in DN1 cells and unswitched memory populations ^6, 21, 22^. As in our ICU cohort, this phenotype highly correlated with disease severity and is preferentially observed in African American patients, a population disproportionally represented in severe COVID-19 in general and in our ICU in particular.

Direct comparison of the CoV-A cluster to patients with active, EF-driven SLE revealed strikingly similar features within the DN compartment (Fig 3c). DN composition, a key indicator of EF response activation, is clearly skewed towards the ASC-associated DN2 subset in the CoV-A group, and was similar to patients with active SLE (Fig 3d-e). Thus, DN2:DN1 ratios, a reflection of EF to follicular response dynamics, are significantly higher in this group (Fig 3f). Of additional interest, the EF-dominated phenotype is associated with a contraction of unswitched memory cells, a feature consistently observed in SLE and other autoimmune diseases ^21, 23, 24^ (Fig 3g). Chemokine receptor analysis of EF populations in patients with COVID-19 shows a decrease in follicular homing predisposition through CXCR5 (Fig 3h). Instead, CXCR3 expression, in conjunction with expression the chemokine CXCL10 (IP-10) is suggestive of peripheral homing to inflamed tissue sites such as the lung as previously described in human influenza infection ^25^ (Fig 3h).

The EF response characterization has identified the pathway as peripherally focused, inflammatory, and highly associated with IL-6 and IP-10 ^22, 26, 27^. Both of these factors have now been identified as biomarkers of poor prognosis in the COVID-19 literature ^28^ and could be identified at increased levels in the CoV-A cluster (Fig 3i-j). In keeping with those studies, the four patients with the highest levels of IL-6 did not survive (Fig 3j). CRP, an acute phase reactant in the IL-6 pathway, has been increasingly used as a biomarker of COVID-19 severity and was also increased in ICU-C (Fig 3k). Expression of IL-6 and IP-10 were both correlated with CRP in patients with COVID-19 (Fig 3l-m). Importantly, DN2 prevalence within the DN compartment was also significantly associated with CRP levels, indicating that the extent of EF activation is a correlate of disease severity (Fig 3n).

The expansion of ASCs in the ICU-C group raises important questions about their function and specificity. Antigen specific responses against the receptor-binding domain (RBD) of the SARS-CoV-2 spike protein were higher in the severe vs. mild groups (Fig 4a). This observation could represent non-neutralizing responses to control the high degree of viral replication, and highlights the possibility of antibody mediated enhancement of disease or pathogenic antibody responses. In turn, pathogenic ASC function could result in autoantibody production^29, 30^ or pro-inflammatory cytokines ^31^.

**Figure 4.**
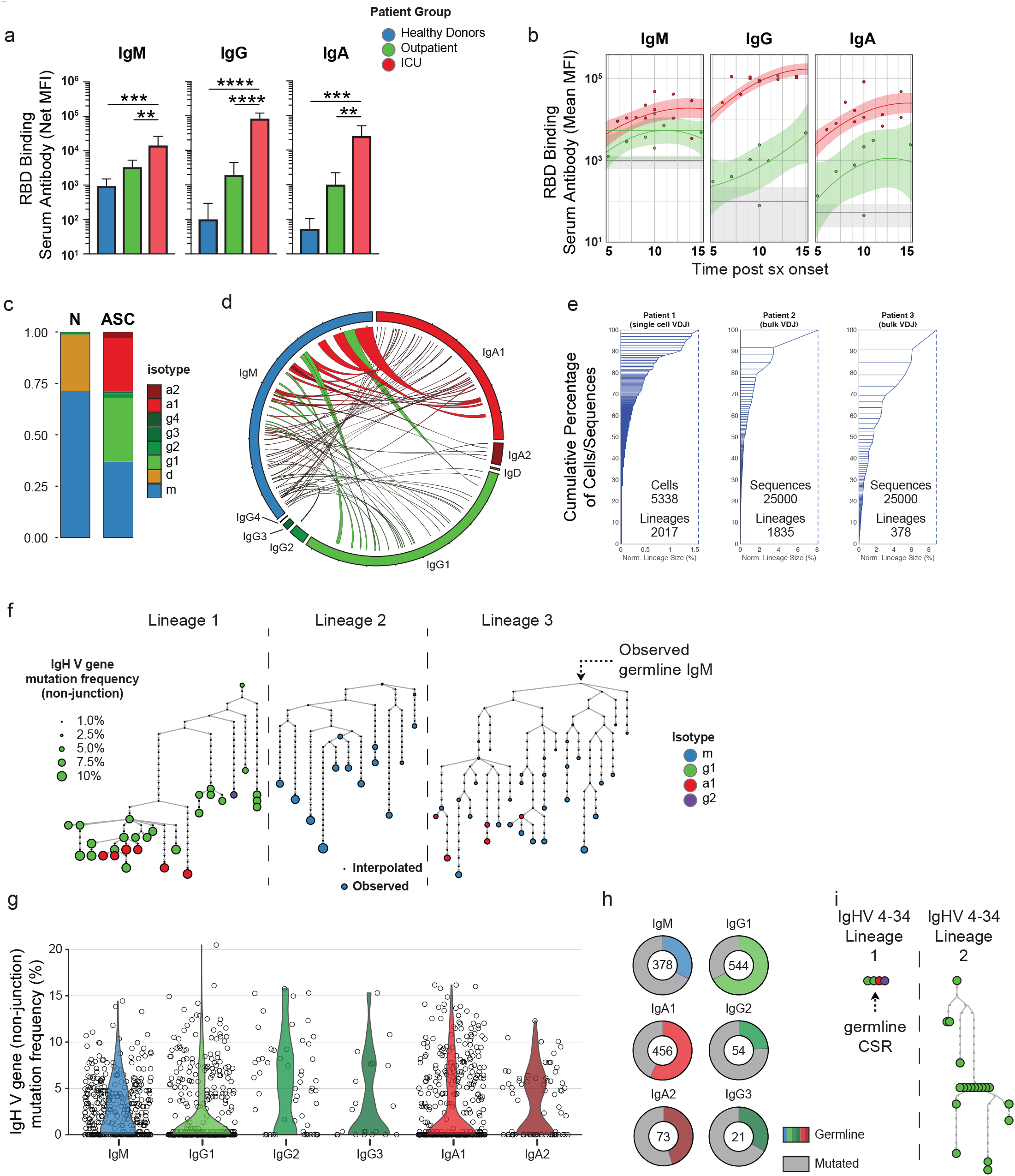
ASCs from ICU patients are antigen specific but display low levels of SHM. (a) Serum RBD-specific antibody responses grouped by indicated isotype in HD, ICU, or outpatient groups. (b) RBD-specific IgM, IgG, and IgA level as a function of time in HD, ICU, or outpatient groups. Solid lines – quadratic regression; Shaded areas – 95% CI of quadratic regression. (c) Isotype usage of single cell VDJ repertoire of sorted ASCs from CoV-A group patient (Patient 1) indicated in Fig 3a. (d) Circos plot of scVDJ repertoire from Patient 1 displaying observed lineages containing cellular members in both unswitched (IgM) and switched compartments. (e) Sample clonality of ASCs. Left, scVDJ from Patient 1. Middle-right, bulk VDJ repertoire sequencing following CD138+ ASC enrichment. (f) Representative lineage trees from scVDJ patient 1 of lineages displaying evidence of somatic hypermutation. (g) Distribution of average lineage mutation frequencies by isotype in scVDJ patient 1 (h) Percentage of lineages in each isotype that display exclusively germline IgHV sequences. (i) Representative lineages utilizing IgHV 4-34 with intact AVY hydrophobic patches in scVDJ patient 1. (a) ANOVA analysis with Tukey’s multiple comparison testing between groups. *p <= 0.05; **p <= 0.01; ***p <= 0.001; **** p <= 0.0001.

This high Ag-specific responsiveness is broad with participation of IgM, IgA, and IgG antibodies (Fig 4a). Importantly, ICU-C responses are higher than outpatient responses and occur very early in the infection course with significantly elevated titers by day 5 post symptom onset (Fig 4b). In accordance with these data, single cell VDJ repertoire sequencing of ASCs from a critically ill patient (Fig 3a) reveal that these responses utilized a balanced IgM, IgG1, and IgA1 signature (Fig 4c-d). Moreover, more than 3% of all of the lineages identified this patient displayed active contemporaneous connection between IgM, and IgG1 or IgA1, thereby demonstrating ongoing isotype switch consistent with guided maturation of the immune response (Fig 4d). Also consistent with ongoing maturation and antigen-selection, the ASC VDJ repertoire was characterized by the presence of clonal expansions with numerous clones contributing more than 0.1% of the entire repertoire and multiple clones representing 1-8% – a finding confirmed by bulk VDJ sequencing of two additional ICU-C patients (Fig 4e). In addition, single cell sequencing identified individual multimember clonal lineages with complex branching patterns, class switching, and broad ranges of somatic hypermutation indicative of robust antigen selection (Fig 4f). Nevertheless, a majority of the lineages identified had remarkably low mutation frequencies – especially lineages comprising the IgA1 and IgG1 compartments (Fig 4g). Indeed, more than half of the lineages obtained expressed germline VH genes. This pattern is consistent with the presence of newly recruited EF clones as previously reported by our group in the SLE EF responses ^21^ (Fig 4h). By analogy with the SLE response where unmutated, switched ASC displayed a high degree of autoreactivity, it is possible that in severe COVID-19 these clones might represent viral responders, a feature that will require further analysis. A hallmark of SLE is defective tolerance resulting in increased levels of disease-specific IgHV 4-34 B cells and autoantibodies with established intrinsic autoreactivity determined by a hydrophobic patch encoded in the germline framework 1^32^. This patch is selected against in healthy individuals to allow the expression of non-autoreactive, protective IgHV 4-34 antibodies ^33^. In contrast, this germline patch could be readily observed in the repertoire dataset with 34 of 40 VH4-34 expressing lineages even in expanded clones (9/34) and in those with significant levels of overall VH mutation (Fig 4h). In all, while the relatively low frequency of IgHV 4-34 clones argues against the expansion of SLE-related autoantibodies in severe COVID-19, the high frequency of IgHV 4-34 retaining germline determinants of autoreactivity could be indicative of tolerance breakdown. Further studies to understand the autoreactivity of COVID-19 ASC will be critical to understand their contribution to the extensive immunopathology now well documented in these patients^34^.

COVID-19 has vexed clinicians across the globe with its spectrum of illness severity and seemingly maladaptive immune response. Through careful humoral immunophenotyping, the current study offers three important observations in severe illness: 1. While ASCs are robustly expanded in serious infection, their presence cannot be considered a correlate of productive or protective immunity. Indeed, the opposite is true. Robust, antigen-specific humoral immunity early in the disease course carries an ominous prognosis in our dataset. 2. Serious COVID-19 illness is highly associated with active extrafollicular B cell responses reminiscent of active SLE and other autoimmune disorders. 3. Due to disease heterogeneity, immunomodulatory therapy, including IL-6 inhibition may have discordant effects on different patient groups. Finally, it will be critical to determine if, as in SLE, pre-disposition towards extrafollicular B-cell activation may be compounding with socio-economic factors to result in the poor infection outcomes observed in the African American population.

## Methods

### Human Subjects

All research was approved by the Emory University Institutional Review Board (Emory IRB numbers IRB00058507, IRB00057983, and IRB00058271) and was performed in accordance with all relevant guidelines and regulations. Written informed consent was obtained from all participants or, if they were unable to provide informed consent, obtained from designated healthcare surrogates. Healthy donors (n = 36) were recruited using promotional materials approved by the Emory University Institutional Review Board. Subjects with COVID-19 (n = 19) were recruited from Emory University Hospital, Emory University Hospital Midtown and Emory St. Joseph’s Hospital, all in Atlanta, GA, USA. All non-healthy donor subjects were diagnosed with COVID-19 by PCR amplification of SARS-CoV-2 viral RNA obtained from nasopharyngeal or oropharyngeal swabs. Subjects with COVID-19 were included in the study if they were 18 to 80 years of age, not immunocompromised, and had not been given oral or intravenous corticosteroids within the preceding 14 days. Peripheral blood was collected in either heparin sodium tubes (PBMCs) (BD Diagnostic Systems) or serum tubes (serum) (BD Diagnostic Systems). Baseline subject demographics are included in Supplemental Table 2. Study data were collected and managed using REDCap electronic data capture tools hosted at Emory University^35^

### Peripheral Blood Mononuclear Cell Isolation and Plasma Collection

Peripheral blood samples were collected in heparin sodium tubes and processed within 6 hours of collection. PBMCs were isolated by density gradient centrifugation at 1000 × g for 10 minutes. Aliquots from the plasma layer were collected and stored at *80C until use. PBMCs were washed 2 times with RPMI at 500 × g for 5 minutes. Viability was assessed using trypan blue exclusion and live cells were counted using an automated hemocytometer.

### Flow cytometry

2 million isolated PBMCs were centrifuged and resuspended in 75ul FACS buffer (PBS + 2% FBS) + 5ul Fc receptor block (Biolegend Cat# 422302) for 5 min RT. For samples stained with anti-IgG, it was observed that Fc block inappropriately interfered with staining, so a pre-incubation step of the anti-IgG alone for 5 min at RT was added prior to the addition of the block. 25ul of Ab cocktail was added (100ul staining reaction), and samples were incubated for 20min at 4 degrees. Cells were washed in PBS, and resuspended in a PBS dilution of Zombie NIR fixable viability dye (Biolegend Cat# 423106). Cells were washed, and fixed at 0.8% PFA for 10 min at RT in the dark before a final wash and resuspension for analysis.

For intracellular staining, the 0.8% PFA fix step was omitted, and an additional wash was added following viability staining. Fixation and permeabilization of the cells was carried out as per manufacturer’s instructions using the True-Nuclear™ Transcription Factor Buffer Set (Biolegend Cat# 424401). Following permeabilization, cells were stained with ICS-directed antibodies diluted in permeabilization buffer as listed.

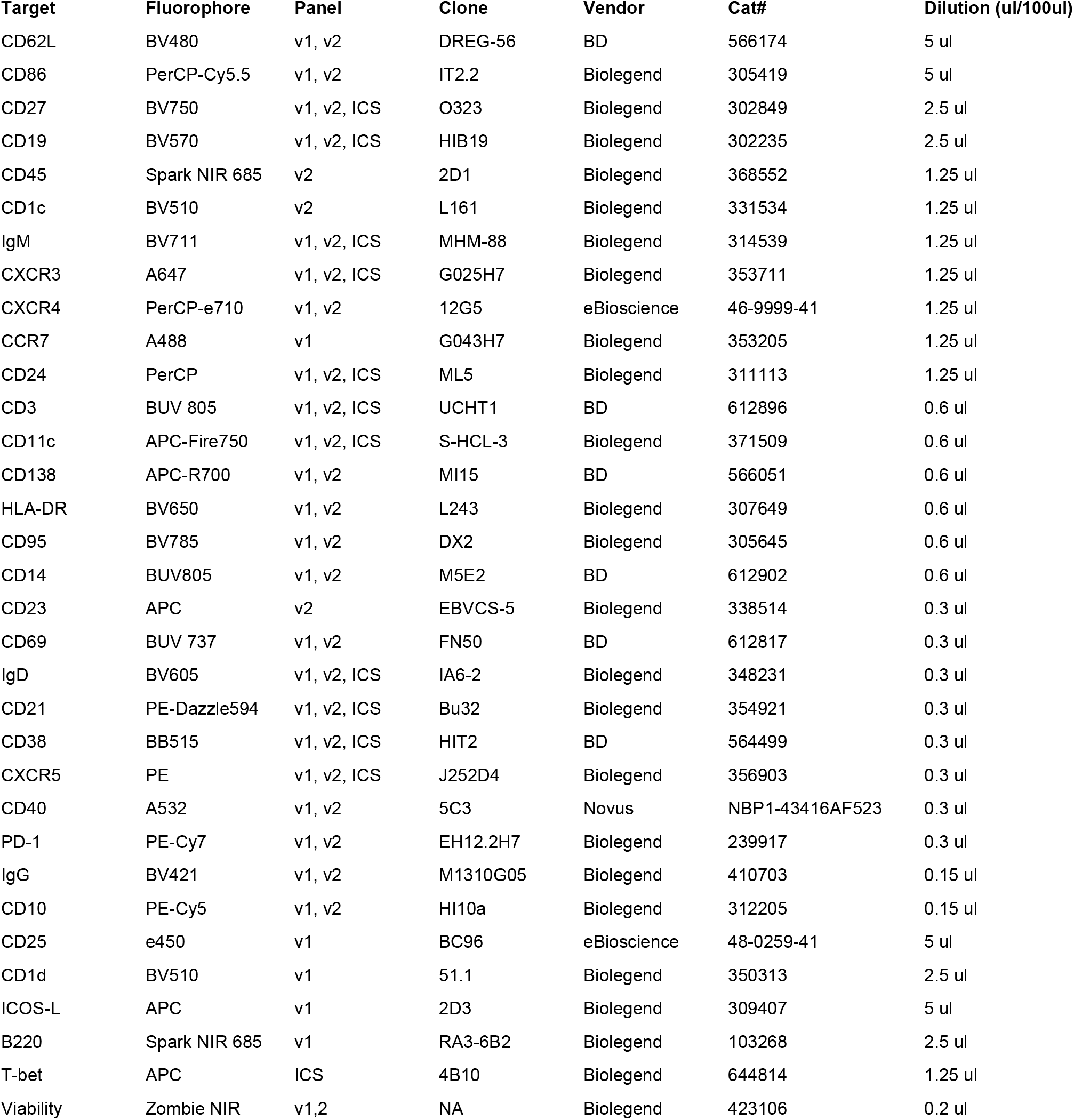

Cells were analyzed on a Cytek Aurora flow cytometer using Cytek SpectroFlo software. Up to 3 × 10^6^ cells were analyzed using FlowJo v10 (Treestar) software. For UMAP projections, all samples stained with panel 2 were downsampled using the DownSample plugin (V3.3) available on the FlowJo Exchange. All samples were concatenated to create a single composite, and a UMAP algorithm for dimensionality reduction was applied using the UMAP plugin (V3.1) available on the FlowJo Exchange. The composite sample was re-gated for all primary and secondary populations to aid in visual overlays in exploration of the UMAP projections.

### Analysis software

Computational analysis was carried out in R, version 3.6.2 (release 12.12.2019). Heatmaps were generated using the pheatmap library (version 1.0.12), with data pre-normalized (log-transformed z-scores calculated per feature) prior to plotting. Clustering was carried out using Ward’s method. Custom plotting such as Ag-specific response curves was performed using the ggplot2 library for base analysis, and then post-processed in Adobe Illustrator. Circos plotting was carried out using Circos software, version 0.69-9. Lineage trees were claculated using GLaMST (Grow Lineages along Minimum Spanning Tree) software^36^, run on matlab, and then visualized in R using the iGraph package, version 1.2.4.2. Exported trees were then post-processed in Adobe Illustrator. Statistical analyses were performed directly in R, or in Graphpad Prism, version 8.2.1.

### Flow Cytometry and Cell Sorting of B Cell Subsets

Frozen cell suspensions were thawed at 37°C in RPMI + 10% FCS and then washed and resuspended in FACS buffer [PBS + 2% FCS]. The cells were incubated with a mix of fluorophore-conjugated antibodies for 30 min on ice. The cells were washed in PBS and then incubated with the Live/Dead Fixable Aqua Dead Cell Stain (Thermofisher) for 10 min at room temperature. After a final wash in FACS buffer, the cells were resuspended in FACS buffer at a concentration of 107 cells/ml for cell sorting on a three-laser BD FACS (BD Biosciences). ASCs were gated as CD3− CD14− CD16− CD19+CD38+CD27+ single live cells whereas naïve B cells were gated as CD3− CD14− CD16− IgD19+CD27-IgD+CD38+ single live cells.

For bulk sequencing preparations, B cells were enriched StemCell’s Human Pan-B Cell Enrichment Kit (Catalog #19554; negative selection of CD2, CD3, CD14, CD16, CD36, CD42b, CD56, CD66b, CD123). CD138+ ASCs were further enriched using CD138+ selection beads according to the manufacturer’s instructions (Miltenyi Biotec, Cat# 130-051-301).

### Library preparation and sequencing

Cells were immediately counted using a hemocytometer and adjusted to 1000 cells/µL to capture 10,000 single cells/sample loaded in the 10x-Genomics Chromium according to the manufacturer’s standard protocol (Chromium Next GEM Single Cell V(D)J Reagent Kits v1.1). The 10x-Genomics v2 libraries were prepared using the 10× Genomics Chromium Single Cell 5′ Library Construction Kit as per the manufacturer’s instructions. Libraries were sequenced, on an Illumina NovaSeq (paired-end; 2X 150 bp; read 1: 26 cycles; i7 index: 8 cycles, i5 index: 0 cycles; read 2: 98 cycles) such that more than 70% saturation can be achieved with sequence depth of 5000 reads per cell.

### Cytokine Immunoassays

Plasma levels of IL-6 was carried out by ELISA using a Human IL-6 Quantikine ELISA Kit according to the manufacturer’s instructions (R+D Biosystems, Cat # HS600C). CRP was measured in a singleplex immunoassay (MilliporeSigma, Burlington, MA) in a Luminex-200 platform following manufacturer’s protocol (25 uL of 1:40,000 dilution in duplicates).

### Carbodiimide coupling of microspheres to *SARS-CoV-2* antigens

Two *SARS-CoV-2* proteins were coupled to MagPlex® Microspheres of different regions (Luminex; Austin, TX, USA). Nucleocapsid (N) protein expressed from *E. coli* (N-terminal His6) was obtained from Raybiotech (230-01104-100) and the Receptor Binding Domain (RBD) of Spike protein expressed from HEK293 was obtained from the laboratory of Dr. Jens Wrammert^37^. Coupling was carried out at room temperature following standard carbodiimide coupling procedures. Concentrations of coupled microspheres were confirmed by Bio-rad T20 Cell Counter.

### Luminex proteomic assays for measurement of anti-antigen antibody

Fifty µL coupled microsphere mix was added to each well of clear bottom 96-well black polystyrene microplates (Greiner Bio-One) at a concentration of 1000 microspheres per region per well. All wash steps and dilutions were accomplished using 1% BSA, 1X PBS assay buffer. Serum was assayed at 1:500 dilution and surveyed for antibodies against N and RBD. After a one-hour incubation in the dark on a plate shaker at 800 rpm, wells were washed five times in 100 µL of assay buffer, using a BioTek 405 TS plate washer, then applied with 3 µg/mL PE-conjugated Goat Anti-Human IgA, IgG and/or IgM (Southern Biotech; Birmingham, AL, USA). After 30 minutes of incubation at 800 rpm in the dark, wells were washed three times in 100 µL of assay buffer, resuspended in 100 µL of assay buffer, and analyzed using a Luminex FLEXMAP 3D® instrument (Luminex; Austin, TX, USA) running xPonent 4.3 software. Median Fluorescent Intensity (MFI) using combined or individual detection antibodies (anti-IgA/anti-IgG/anti-IgM) was measured using the Luminex xPONENT software. The background value of assay buffer was subtracted from each serum sample result to obtain Median Fluorescent Intensity minus Background (MFI-B; Net MFI).

### Statistical Analysis

Statistical analysis was carried out using Prism statistical analysis software. For each experiment, the type of statistical testing, summary statistics, and levels of significance can be found in the figures and corresponding figure legends

## Data Availability

Relevant human clinical data from the manuscript will be shared as required by the NIH upon formal acceptance to a peer-reviewed journal.

## Acknowledgements

We would like to thank all of the healthy volunteers, patients, and their families for their selfless participation in this study. We would also like to thank the nurses, staff, and providers in the 71 ICU in Emory University Hospital Midtown and the 2E ICU in Emory Saint Joseph’s Hospital without whom our work could not have been possible. We would like to acknowledge the contributions of Dr. Bashar Staitieh, Dr. David Murphy, Dr. William Bender, Dr. Colin Swenson, Dr. Michael Horwath, Dr. David Alter, Sang Le, Dr. Vanessa Engineer, Mindy Hernandez, and John Varghese for their time and expertise. We would like to thank Dr. Jens Wrammert and Dr. Florian Krammer for the COVID-19 antigens. Extensive efforts by the Pediatric/Winship Flow Cytometry Core, and the Flow Cytometry Core at the Emory Vaccine Center made this work possible.

## COIs

Dr. Lee is the founder of MicroB-plex, Inc and has research grants with Genentech. Dr. Hu has consulted for ViveBio LLC, AARP Inc, and Biogen Inc; has received research support from Fujirebio US; and has a patent on CSF-based diagnosis of FTLD-TDP.

---

**Supplementary Figure 1.**
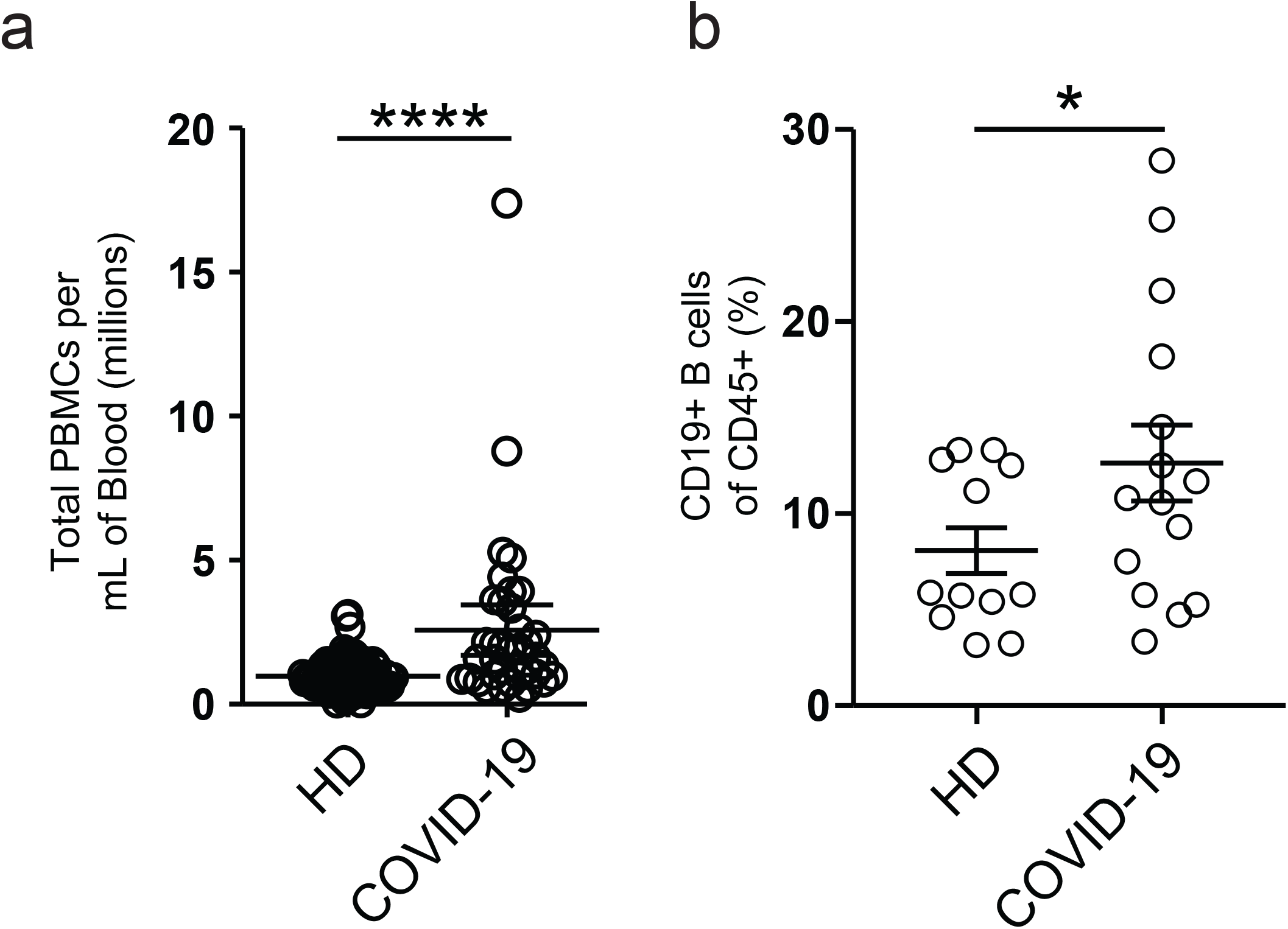
Cellularity of COvID-19 patient blood samples. (a) Calculated total PBMC yield per mL of patient blood from HD, or donors with COVID-19. (b) Frequency of CD19+ B cells of CD45+ cells in HD vs donors with COVID-19

**Supplementary Figure 2.**
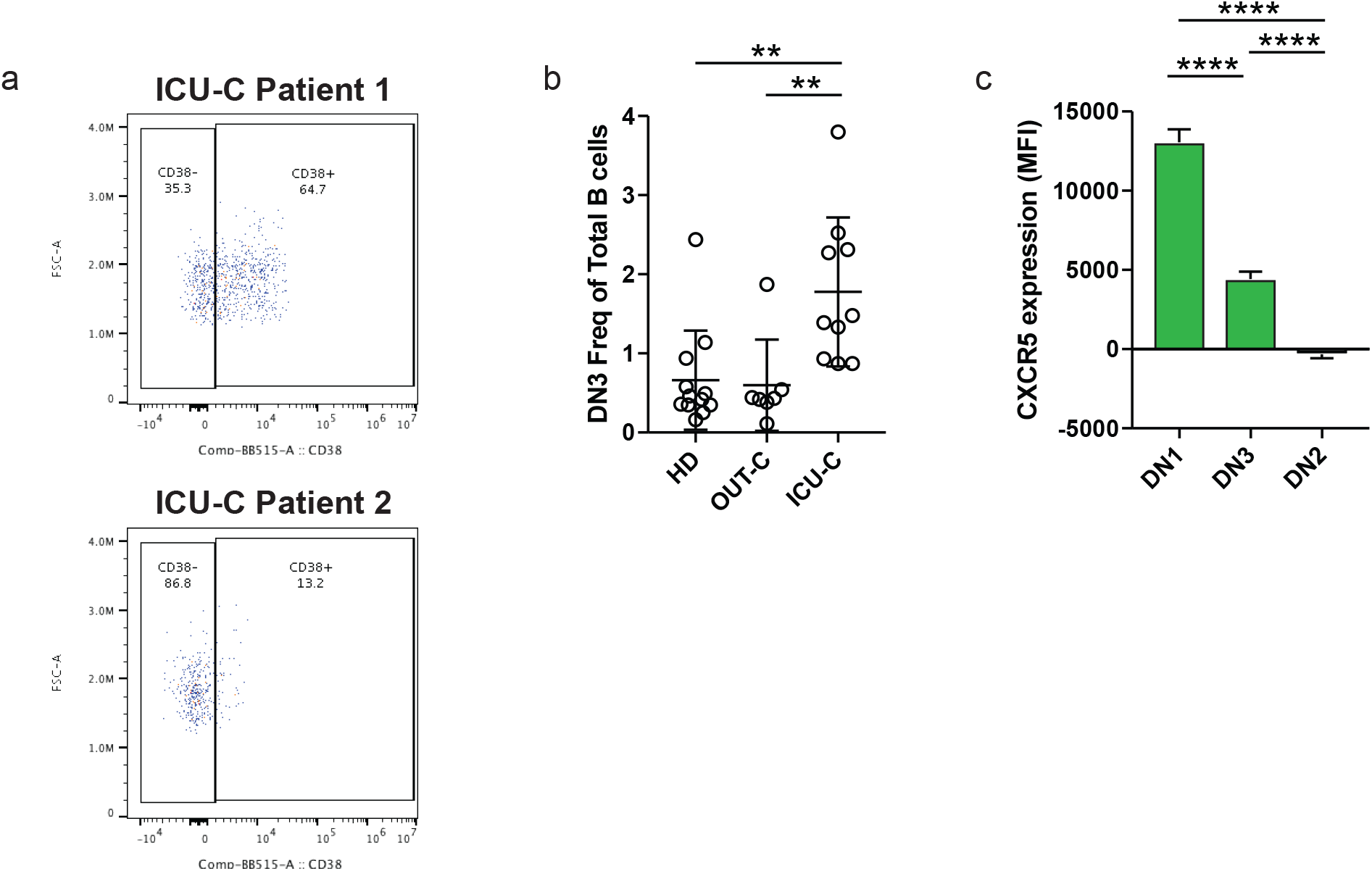
DN3 cells are expanded in the ICU-C cohort. (a) CD38 expression by DN3 cells in two ICU-C patients. (b) DN3 frequency of total CD19+ B cells in HD, OUT-C, and ICU-C cohorts. (c) CXCR5 expression (geometric MFI) of DN1, DN2, and DN3 populations across all patient samples.

**Supplementary Figure 3.**
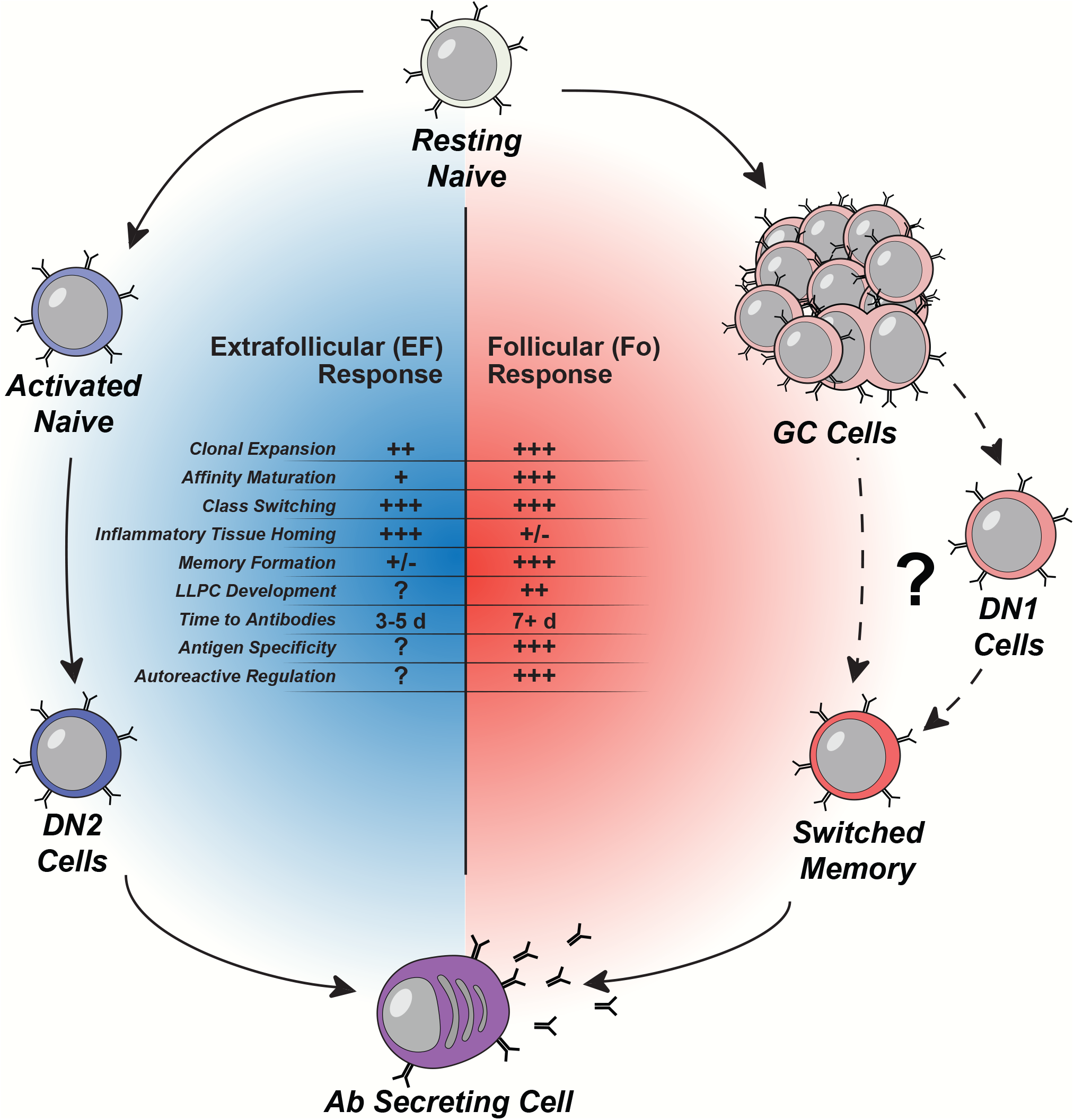
Comparison of follicular and extrafollicular response. Illustration of the recognized cellular populations, and functional properties of follicular versus extrafollicular responses

**Table S1.**
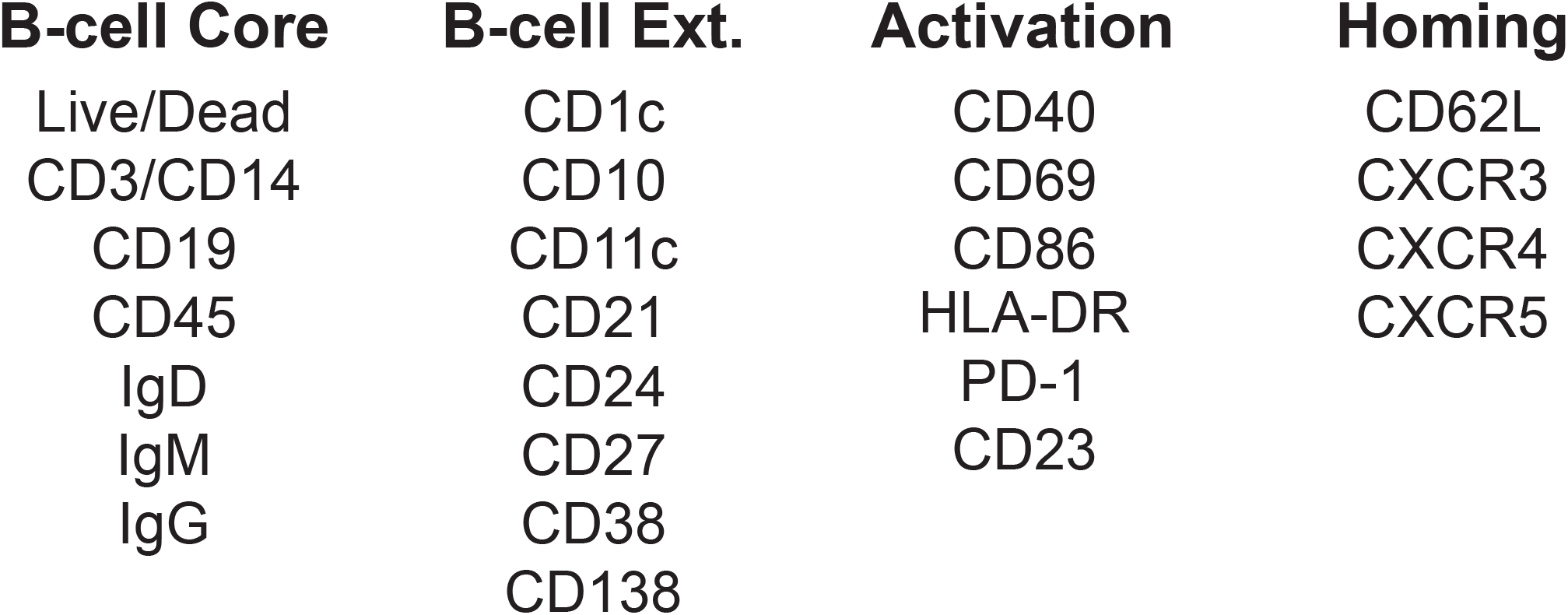
Spectral Flow B cell Panel Design. Standardized panel design for the broad characterization and study of peripheral blood B cells

**Table S2.**
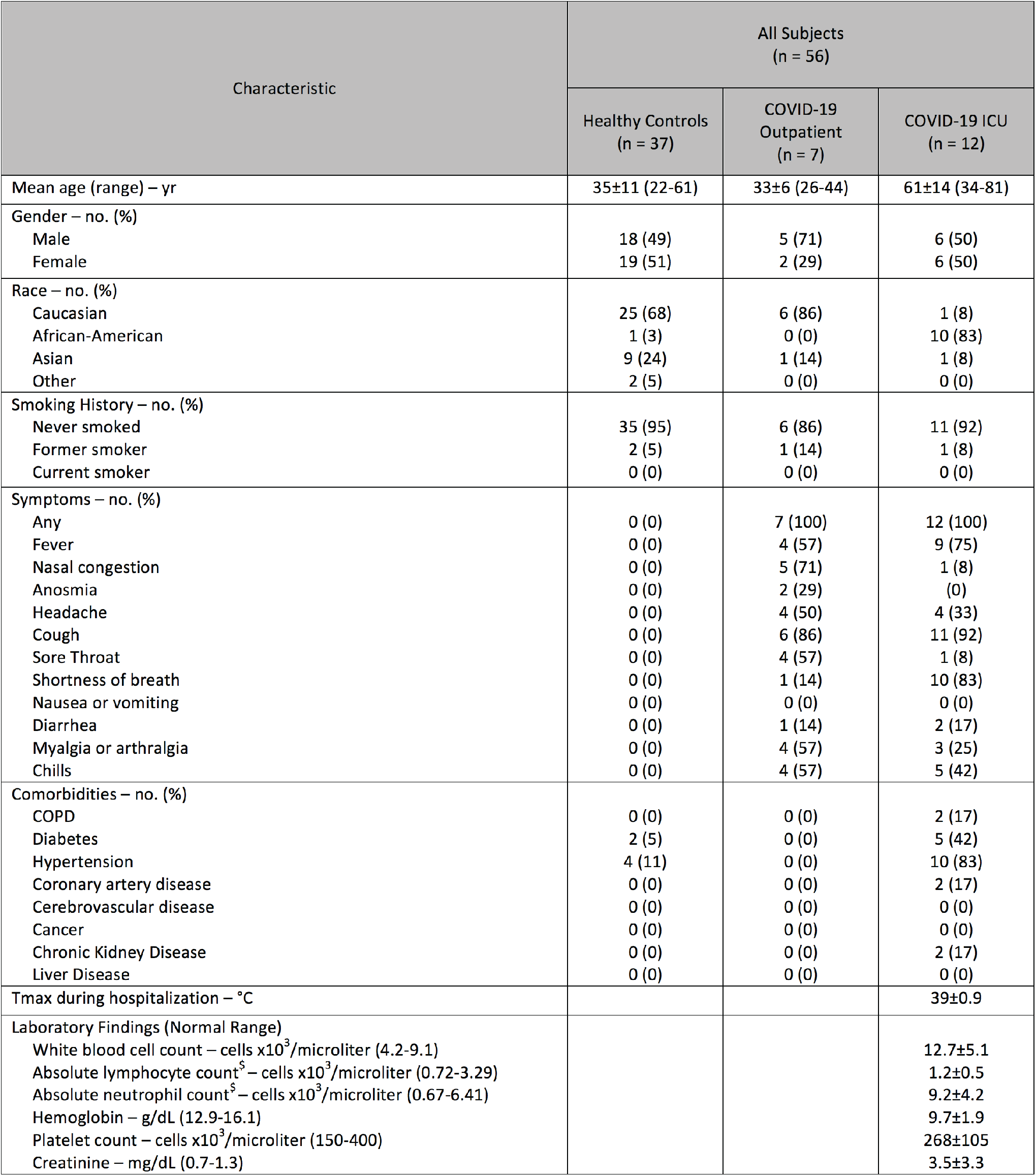
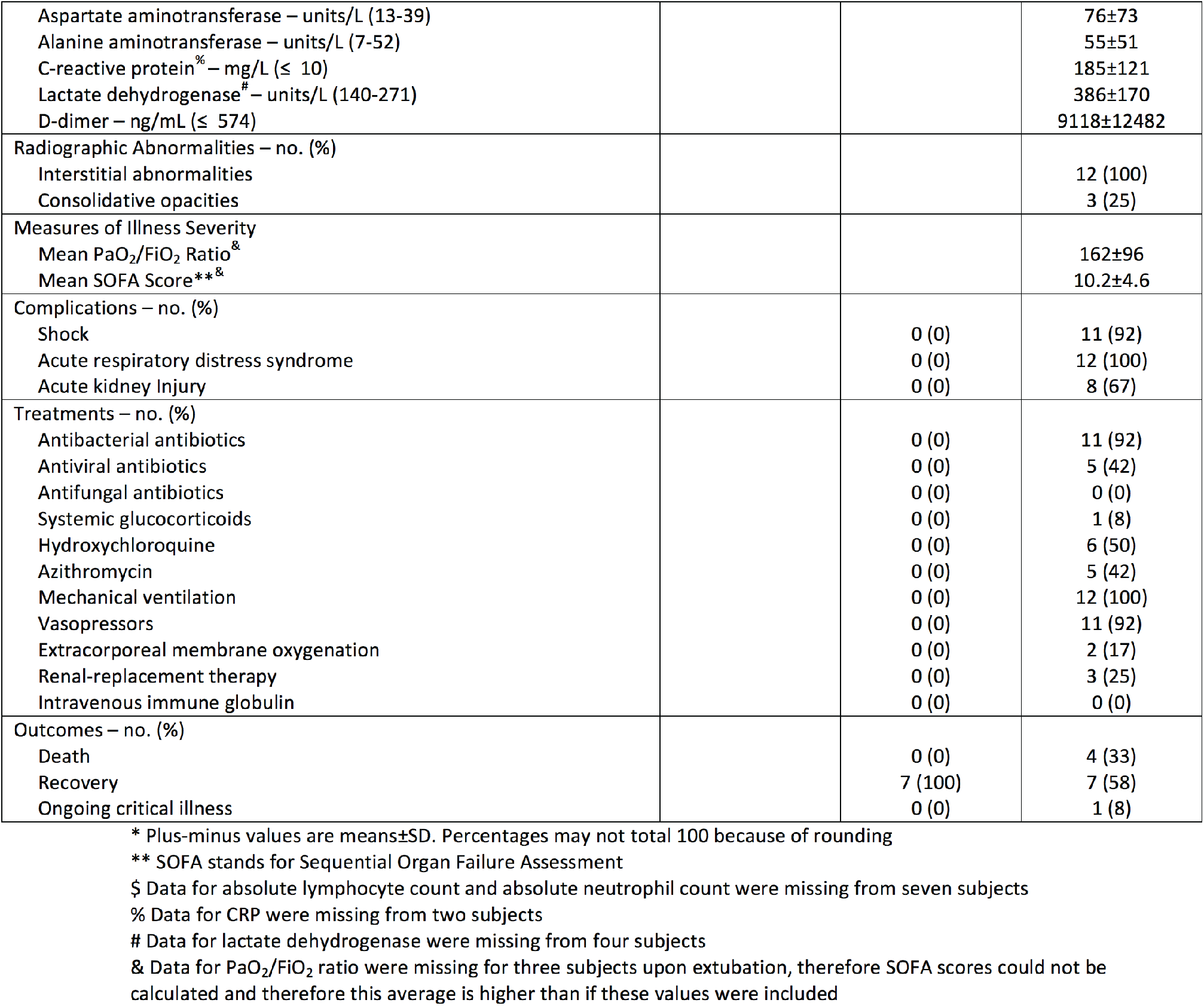
Baseline Demographic Characteristics of Research Subjects*.

| Characteristic | All Subjects<br>(n = 56) |  |  |
| --- | --- | --- | --- |
|  | Healthy Controls<br>(n = 37) | COVID-19<br>Outpatient<br>(n = 7) | COVID-19 ICU<br>(n = 12) |
| Mean age (range) – yr | 35±11 (22-61) | 33±6 (26-44) | 61±14 (34-81) |
| Gender – no. (%) |  |  |  |
| Male | 18 (49) | 5 (71) | 6 (50) |
| Female | 19 (51) | 2 (29) | 6 (50) |
| Race – no. (%) |  |  |  |
| Caucasian | 25 (68) | 6 (86) | 1 (8) |
| African-American | 1 (3) | 0 (0) | 10 (83) |
| Asian | 9 (24) | 1 (14) | 1 (8) |
| Other | 2 (5) | 0 (0) | 0 (0) |
| Smoking History – no. (%) |  |  |  |
| Never smoked | 35 (95) | 6 (86) | 11 (92) |
| Former smoker | 2 (5) | 1 (14) | 1 (8) |
| Current smoker | 0 (0) | 0 (0) | 0 (0) |
| Symptoms – no. (%) |  |  |  |
| Any | 0 (0) | 7 (100) | 12 (100) |
| Fever | 0 (0) | 4 (57) | 9 (75) |
| Nasal congestion | 0 (0) | 5 (71) | 1 (8) |
| Anosmia | 0 (0) | 2 (29) | 0 (0) |
| Headache | 0 (0) | 4 (50) | 4 (33) |
| Cough | 0 (0) | 6 (86) | 11 (92) |
| Sore Throat | 0 (0) | 4 (57) | 1 (8) |
| Shortness of breath | 0 (0) | 1 (14) | 10 (83) |
| Nausea or vomiting | 0 (0) | 0 (0) | 0 (0) |
| Diarrhea | 0 (0) | 1 (14) | 2 (17) |
| Myalgia or arthralgia | 0 (0) | 4 (57) | 3 (25) |
| Chills | 0 (0) | 4 (57) | 5 (42) |
| Comorbidities – no. (%) |  |  |  |
| COPD | 0 (0) | 0 (0) | 2 (17) |
| Diabetes | 2 (5) | 0 (0) | 5 (42) |
| Hypertension | 4 (11) | 0 (0) | 10 (83) |
| Coronary artery disease | 0 (0) | 0 (0) | 2 (17) |
| Cerebrovascular disease | 0 (0) | 0 (0) | 0 (0) |
| Cancer | 0 (0) | 0 (0) | 0 (0) |
| Chronic Kidney Disease | 0 (0) | 0 (0) | 2 (17) |
| Liver Disease | 0 (0) | 0 (0) | 0 (0) |
| Tmax during hospitalization – °C |  |  | 39±0.9 |
| Laboratory Findings (Normal Range) |  |  |  |
| White blood cell count – cells x10<sup>3</sup>/microliter (4.2-9.1) |  |  | 12.7±5.1 |
| Absolute lymphocyte count<sup>s</sup> – cells x10<sup>3</sup>/microliter (0.72-3.29) |  |  | 1.2±0.5 |
| Absolute neutrophil count<sup>s</sup> – cells x10<sup>3</sup>/microliter (0.67-6.41) |  |  | 9.2±4.2 |
| Hemoglobin – g/dL (12.9-16.1) |  |  | 9.7±1.9 |
| Platelet count – cells x10<sup>3</sup>/microliter (150-400) |  |  | 268±105 |
| Creatinine – mg/dL (0.7-1.3) |  |  | 3.5±3.3 |
| Aspartate aminotransferase – units/L (13-39) |  |  | 76±73 |
| Alanine aminotransferase – units/L (7-52) |  |  | 55±51 |
| C-reactive protein<sup>%</sup> – mg/L (≤ 10) |  |  | 185±121 |
| Lactate dehydrogenase<sup>#</sup> – units/L (140-271) |  |  | 386±170 |
| D-dimer – ng/mL (≤ 574) |  |  | 9118±12482 |
| Radiographic Abnormalities – no. (%) |  |  |  |
| Interstitial abnormalities |  |  | 12 (100) |
| Consolidative opacities |  |  | 3 (25) |
| Measures of Illness Severity |  |  |  |
| Mean PaO<sub>2</sub>/FiO<sub>2</sub> Ratio<sup>&</sup> |  |  | 162±96 |
| Mean SOFA Score<sup>**&</sup> |  |  | 10.2±4.6 |
| Complications – no. (%) |  |  |  |
| Shock |  | 0 (0) | 11 (92) |
| Acute respiratory distress syndrome |  | 0 (0) | 12 (100) |
| Acute kidney Injury |  | 0 (0) | 8 (67) |
| Treatments – no. (%) |  |  |  |
| Antibacterial antibiotics |  | 0 (0) | 11 (92) |
| Antiviral antibiotics |  | 0 (0) | 5 (42) |
| Antifungal antibiotics |  | 0 (0) | 0 (0) |
| Systemic glucocorticoids |  | 0 (0) | 1 (8) |
| Hydroxychloroquine |  | 0 (0) | 6 (50) |
| Azithromycin |  | 0 (0) | 5 (42) |
| Mechanical ventilation |  | 0 (0) | 12 (100) |
| Vasopressors |  | 0 (0) | 11 (92) |
| Extracorporeal membrane oxygenation |  | 0 (0) | 2 (17) |
| Renal-replacement therapy |  | 0 (0) | 3 (25) |
| Intravenous immune globulin |  | 0 (0) | 0 (0) |
| Outcomes – no. (%) |  |  |  |
| Death |  | 0 (0) | 4 (33) |
| Recovery |  | 7 (100) | 7 (58) |
| Ongoing critical illness |  | 0 (0) | 1 (8) |

\* Plus-minus values are means±SD. Percentages may not total 100 because of rounding
\*\* SOFA stands for Sequential Organ Failure Assessment
\$ Data for absolute lymphocyte count and absolute neutrophil count were missing from seven subjects
% Data for CRP were missing from two subjects

**Table S3.**
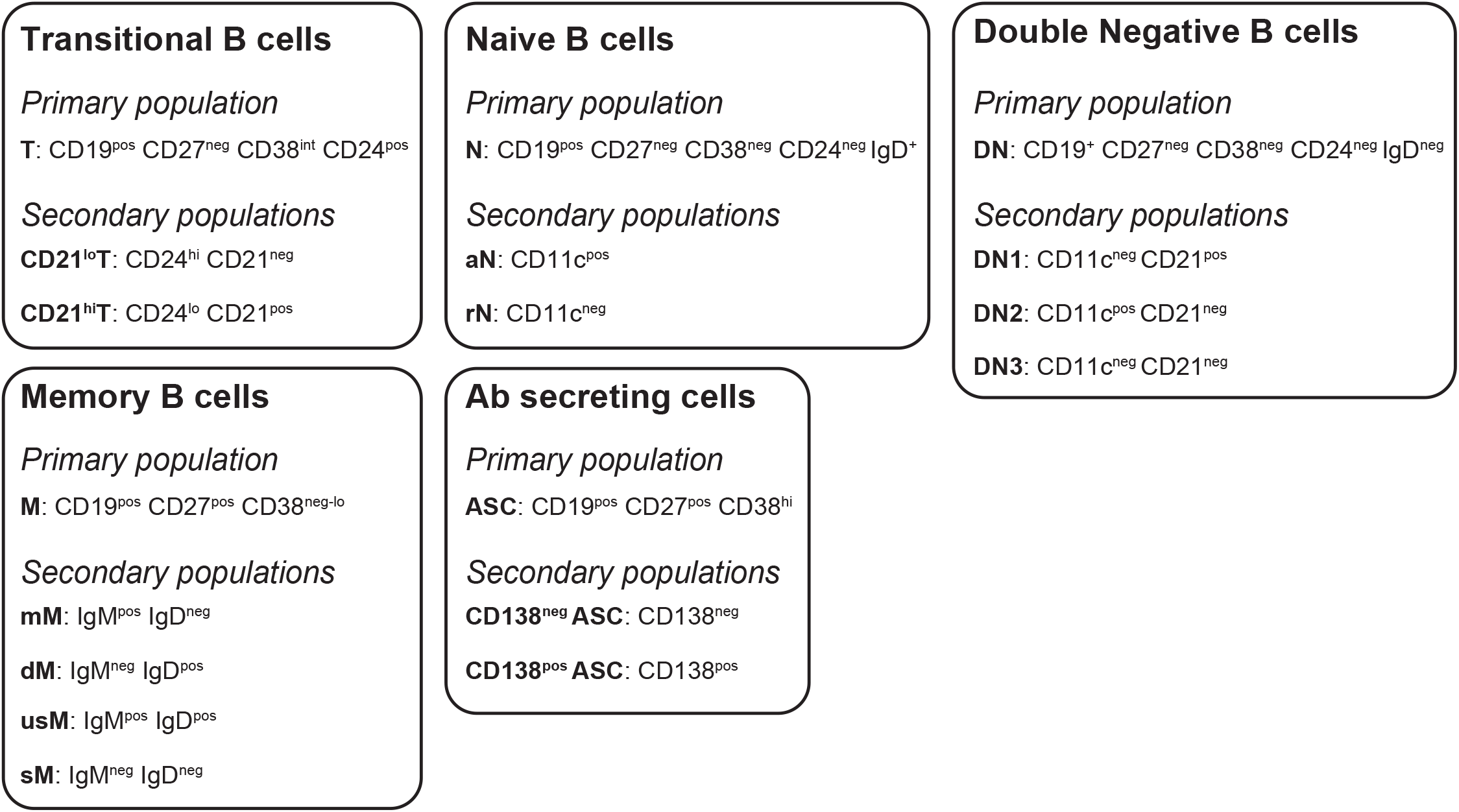
Standard B cell definitions. Surface staining definintions of primary and secondary populations used for multivariate clustering and analysis.

